# Efficacy of virtual reality treatment of phantom leg pain: Results of a randomized clinical trial

**DOI:** 10.64898/2026.04.20.26350810

**Authors:** Elisabetta Ambron, Rand Williamson, Jing-Sheng Li, Maxim Karrenbach, Eric Rombokas, H. Branch Coslett, Laurel J. Buxbaum

## Abstract

Approximately 90% of individuals with limb amputation experience persistent sensation of the missing extremity and up to 85% experience pain in the missing limb, (phantom limb pain; PLP). In this registered clinical trial (NCT05296265), we tested the efficacy of two Virtual Reality (VR) treatments of phantom leg pain. Adaptive randomization was used to assign 36 participants (19 transfemoral, 17 transtibial) to eight sessions of an active or distractor VR treatment. The active VR treatment required leg movements and provided virtual visual feedback. The distractor treatment was a commercial VR treatment for pain. The primary outcome measures were the comparison of pain at baseline versus immediately post-treatment and at 1-week and 8-week follow-up; pain was assessed on two measures from the McGill Pain Questionnaire. The secondary outcome measure, obtained in each session, was average pain intensity since the last treatment. Pain on both primary outcome measures was significantly reduced with moderate effect sizes for the active treatment only; effects on intensity persisted at 1-week follow-up, and effects on pain quality persisted at 8-weeks follow-up. Ratings of pain intensity since the last treatment showed a large effect size for the active treatment and was significant for both treatments. Statistical superiority of the active treatment on the primary and secondary outcomes was not established, but the active treatment showed significant effects on a numerically wider range of measures overall. This is the first clinical trial to show significant efficacy of VR treatment of PLP in a randomized sample of transfemoral and transtibial amputees.

## 1. INTRODUCTION

A common consequence of limb amputation is the sensation that the missing limb remains present, a phenomenon known as *phantom limb* (PL) [56]. In approximately 45–85% of patients with PL [28], these sensations are accompanied by pain, called *phantom limb pain* (PLP). PLP is severe and chronic in 5-10% of patients, causing significant distress and reducing quality of life [7,28].

One theory is that PLP arises from the formation of neuromas—painful growths of nerve tissue at the amputation site [14]. Treatments of neuroma-related pain [4,58] have demonstrated significant pain reduction in patients with identifiable neuromas [58] but are not relevant to the approximately 95% of individuals with PLP for whom neuromas are not identified [41]. A second group of theories attributes PLP to changes in the brain. According to the maladaptive plasticity hypothesis, the loss of sensory input following amputation (deafferentation) disrupts cortical organization in the motor and somatosensory cortices [16] or alters functional connectivity across networks [29,31,38,42]. A variety of treatments including transcranial magnetic stimulation (TMS; [36]) and motor imagery [4,35] have targeted the putative brain mechanisms underlying PLP. Mirror therapy [5,30] appears to be the most commonly employed treatment. In this intervention, a mirror is positioned along the body midline so that the reflection of movements of the intact limb appear to be generated by the missing limb. This method is based on the hypothesis that PLP is due to a mismatch between intact motor commands and the absence of visual and proprioceptive feedback from the missing limb [45,17,56].

Advances in technology have enabled development of virtual reality (VR) and augmented reality (AR) therapies, which extend the principles of mirror therapy by digitally recreating the missing limb. Although a recent meta-analysis of 15 studies showed that mirror therapy and VR treatment are of equivalent efficacy [44,54], VR-based interventions have gained popularity because they offer immersive, interactive environments that increase patient engagement and motivation [3]. Although prior VR interventions have shown promise, a crucial limitation of the literature is that the sample sizes have been small (1-20 participants, see [54]), studies were underpowered and, at least in some instances, the verisimilitude obtainable in earlier virtual reality studies did not match what is readily obtainable today [2,11].

The present registered, randomized clinical trial builds on a prior small clinical trial evaluating the efficacy of both distractor-based and “active” virtual reality (VR) treatments in 7 individuals with transtibial amputation [2–3]. The current study design was novel in the following respects: (i) we used the same VR apparatus in individuals with both transtibial and transfemoral amputation for the first time.; ii) our conservative approach compared an active VR treatment with another VR treatment with established efficacy in treating chronic pain rather than to an inert placebo condition; (iii) to avoid carryover effects between treatments that could occur with a cross-over design, we used stratified randomized assignment to allocate participants to a single treatment; (iv) the active VR treatment was implemented with affordable, readily-available equipment, including motion tracking via Meta Quest 2 controllers rather than laboratory marker-based or markerless motion tracking systems.

## 2. METHODS

### 2.1. Participants

This study was pre-registered at clinicaltrials.gov (NCT05296265) and conducted in accordance with the registered protocol and the guidelines of the Institutional Review Board (IRB) of Thomas Jefferson University. All participants were compensated for their participation and reimbursed for travel expenses. Participants were recruited over a period of three years at Thomas Jefferson University, University of Pennsylvania, and University of Washington (see Figure 1) via support groups for amputees, targeted emails, referrals, and telephone contact. Enrolled individuals were allocated to one of two study arms (active or distractor treatment) using covariate adaptive randomization [51] implemented with R’s carat package. Variables used in the randomization algorithm were average pain severity over the last week, limb telescoping (i.e., sensation of limb shortening; present/absent), level of amputation (Transfemoral/Above Knee Amputation or Transtibial/Below Knee Amputation), and prosthesis use (regular, intermittent/occasional, none). Based on effect sizes in our previous study [2], g*power determined that a sample size of 36 participants was adequate to detect a moderate effect size of.64 with a statistical power of 80% (p<0.05).

**Figure 1:**
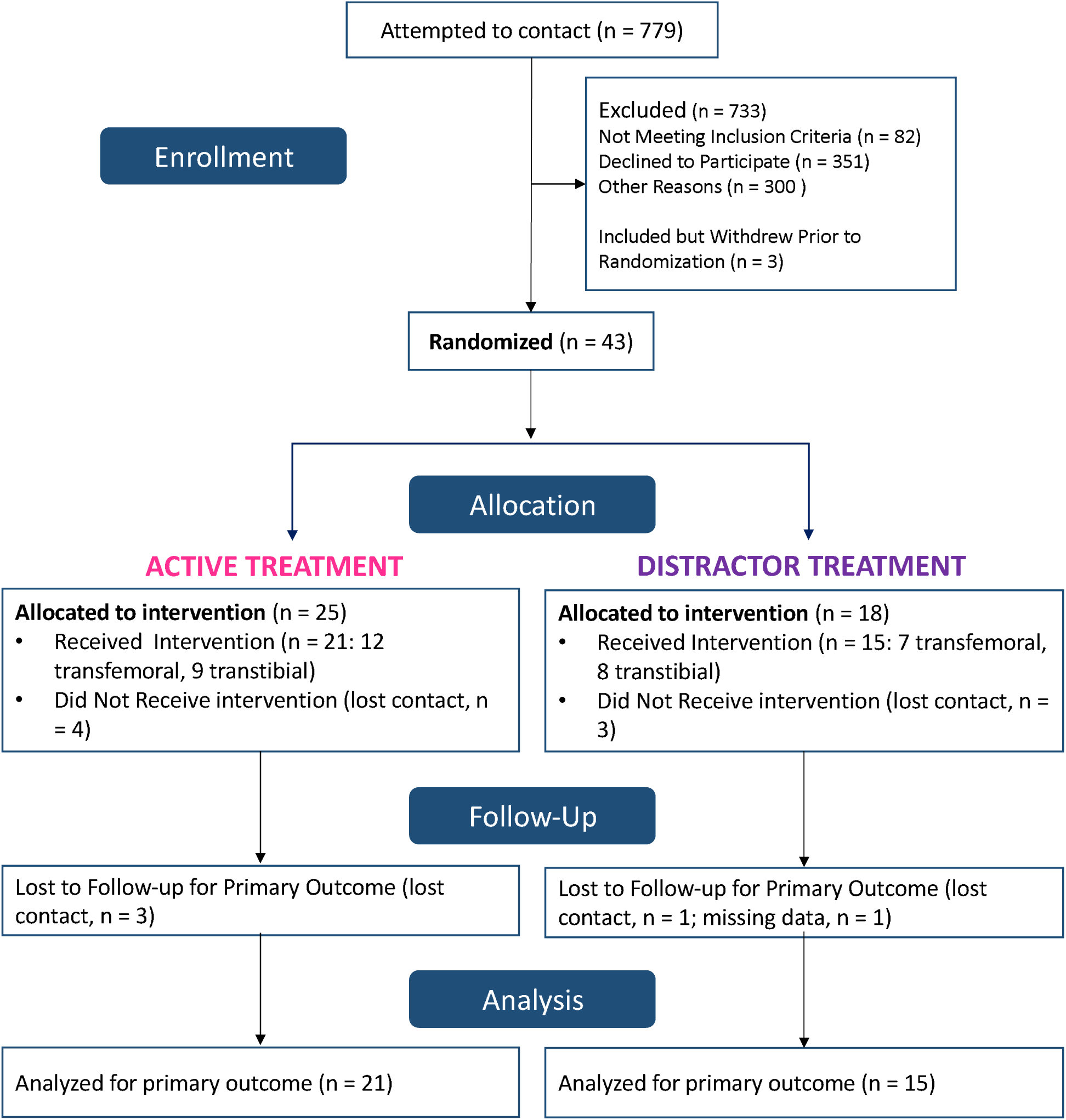
CONSORT diagram of study recruitment pipeline. Highlighted are the final samples of participants in the active and distractor treatment arms, respectively.

Study inclusion criteria were: (1) age between 18 and 100; (2) capacity to provide informed consent; (3) unilateral transfemoral or transtibial amputation > 3 months prior to enrollment; (4) significant cognitive impairment, operationally defined as a Montreal Cognitive Assessment score <17 [33,37]; (5) average PLP > 4 over the preceding 1 month on a scale of 0-10. Specific exclusion criteria were: (1) history of significant medical or neurological disorder such as stroke or moderate to severe traumatic brain injury; (2) history of significant or poorly controlled psychiatric disorders; (3) current significant depression or anxiety; (4) current abuse of alcohol or drugs, prescription or otherwise.

Forty-six people were enrolled in the study; three individuals withdrew prior to randomization and seven withdrew during the study (see Figure 1). The final sample consisted of 36 individuals who completed the treatment phase: 21 individuals completed the active (12 transfemoral, 9 transtibial; 10 female, 11 male; Mean age=59; SE=13.5) and 15 the distractor treatment (7 transfemoral, 8 transtibial; 3 female, 12 male; Mean age=59; SE=10.7).

Study team members who enrolled participants and assigned participants to an intervention had access to the random allocation sequence. The study team was not blinded after assignment to interventions. Participants were informed that the study tested the efficacy of two VR treatments but were not provided information regarding the hypothesized efficacy of one treatment over the other, and care was taken to maintain equipoise throughout.

### 2.2. Overview of procedure

Enrolled participants completed an initial testing battery including the primary outcome measures (described below), followed by a course of Active or Distractor VR treatment as assigned. Both treatments consisted of eight twice-weekly, in-person sessions of approximately 1-hour duration in laboratory spaces at Jefferson Moss Rehabilitation Research Institute, the University of Pennsylvania, or the University of Washington. One and eight weeks following the last treatment session, participants returned for repeat administration of the test battery.

The treatment was delivered in person by a research team member who was trained on a detailed written protocol for administering the treatment and collecting data. Participants were instructed to continue with their usual pain management treatment throughout the course of the project but not initiate a new therapy except the treatment under investigation.

### 2.3. Primary and secondary Outcome Measures

The primary outcome measures were the Pain Intensity and the Pain Rating Index from the McGill Pain Questionnaire, Short Form [32]. The Pain Intensity measure is based on a visual analog scale measuring current pain. The Pain Rating Index is a qualitative measure of pain consisting of a series of 4-point scales assessing 15 characteristics of pain (e.g., throbbing, shooting, cramping, etc.). The measures were administered at baseline, immediately post-treatment, and at 1-week and 8-week follow-up. Pain reductions of at least 2 points has been denoted as clinically meaningful [60].

The secondary outcome measure, obtained in each session, was average pain intensity since the last treatment. In all treatment sessions except the first, participants were asked to rate their average pain since the previous treatment on an 11-point numerical rating pain scale; (0 - minimum score/no pain; 10 maximum score/pain as bad as participants can imagine). For the first session, participants were asked to rate their average pain intensity since the initial testing battery was performed.

### 2.4. Repeated Test Battery

A battery of questionnaires was presented at baseline, immediately after the completion of the treatment, and 1 and 8 weeks after completion of the last treatment. The battery included the primary outcome measures as well as the following additional measures:

1. Frenchay Activities Index (FAI) [24]: a scale measuring physical function and daily activity, used to functionally assess lower limb amputees [62].
2. Twelve-Item Short Form Health Survey (SF-12) [55; 62]: a brief measure of quality of life and functional capacity, used to functionally assess lower limb amputees [62].
3. Pain Interference Scale from the Brief Pain Inventory [10]: an index validated in lower limb amputees [63,64] that assesses the degree to which pain interferes with daily activities using a 0-10 numeric rating scale.
4. Modified Limb Deficiency and Phantom Limb Questionnaire [21]: a questionnaire assessing prosthesis type and usage and non-painful phantom limb experiences, including perceived position (e.g., telescoping) and ability to move the phantom.
5. Hospital Anxiety and Depression Scale (HADS) [59]: a 14-item measure assessing depression and anxiety symptoms.
6. Thirteen-item Pain Catastrophizing Scale [50]: a measure of pain catastrophizing, which has been highly associated with pain severity and disability after amputation [23,26,57].
7. Insomnia Severity Index [6]: a 7-point scale that measures sleep disturbances.

### 2.5. Within-session assessments

As noted, at the beginning of each session we administered the secondary outcome measure, average pain since the last treatment, based on the 11-point scale in the Brief Pain Inventory [10].

### 2.6. VR Usability and Presence Assessments

At baseline we administered the Technology Acceptance scale [34] to assess perceptions of VR. At baseline and after the last treatment session, participants completed the Simulator Sickness Questionnaire [27] and the Brief Slater-Usoh-Steed Presence [48,49,53]; the former assesses cybersickness and the latter assesses the experience of “presence” during the treatment session. The usability of the games was assessed using the System Usability Scale [9] after the last treatment session.

### 2.7. Virtual Reality Treatments

#### 2.7.1. Active treatment

VR System. The active treatment used a Meta Quest 2 (Meta, Menlo Park, CA) head-mounted display and two associated Meta Quest controllers. One controller was secured on the participant’s residual limb (either thigh stump for transfemoral or upper shin/calf for transtibial amputations) with flexible tape and used to control the avatar leg, while the other controller was hand-held for selection and supportive features during the games. For transfemoral subjects, knee/ankle movements (flexion and extension) were imputed based on real-time information from the movements of the hip. For transtibial subjects, the controller provided real-time information to generate virtual movements of the avatar. Games. Subjects assigned to the Active VR treatment participated in custom-made games in which participants were able to see virtual rendering of both legs intact. The motion of the Meta Quest 2 controller attached to the residual limb provided spatial information for the rendering of their virtual leg while playing the games. Participants engaged in two games:

1) Kick: participants kicked a virtual ball using the virtual rendering of the amputated leg to displace objects from a platform, gaining points and bonus kicks at different levels of difficulty (see Figure 2).
2) Quest for Fire: seated on a rolling chair, participants used the virtual leg to move across mazes of different complexities, push obstructions out of the path, and reach a goal ([2]; see Figure 2).

**Figure 2:**
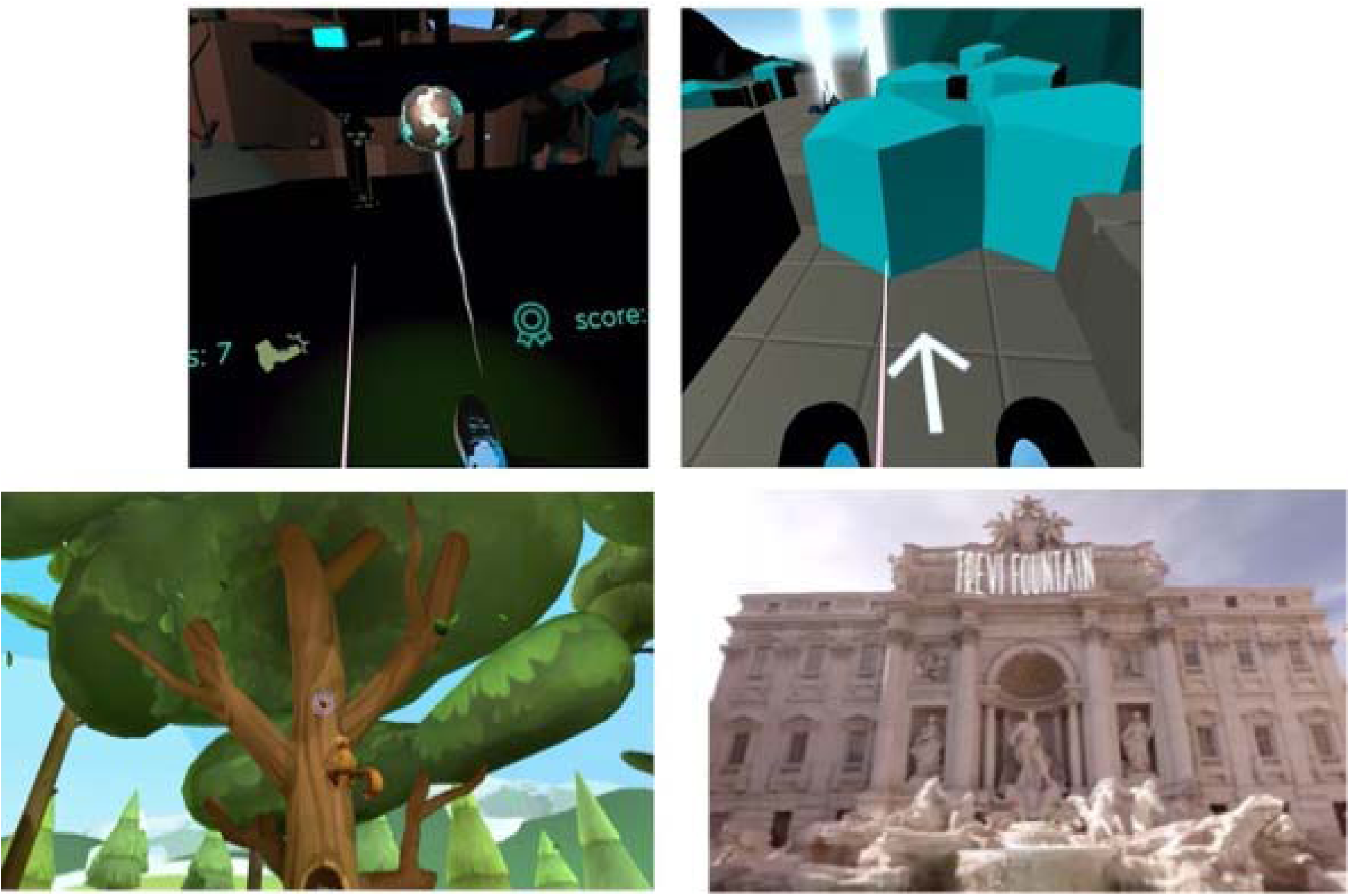
Top Panel: Illustrations of active treatment. Left: Screenshot from Kick VR game; Right: Screenshot from Quest for Fire VR game. Bottom panel: Illustrations of distractor treatment. Left: Screenshot from Serene Lake environment; Right: Screenshot from World Traveler environment.

In each session, participants had flexibility in choosing which game to play.

#### 2.7.2. Distractor treatment

Subjects assigned to the Distractor VR treatment participated in the REAL i-Series® (Penumbra, Inc.) immersive VR experience, which has been demonstrated to reduce pain [8] but lacked the hypothesized “active ingredients” of our Active VR treatment (visual feedback of movement of an extrapolated amputated limb). Wearing a proprietary headset developed by Penumbra, participants navigated through pleasant VR environments without seeing a rendering of their body or moving their legs (see Figure 2).

### 2.8. Statistical Analysis

We performed permutation analyses to test if each treatment individually had a significant effect on the outcome measures and if there was a significant difference between the two treatments. For all primary and secondary outcome measures, we adjusted the significance threshold of p <.05 for multiple comparisons using a False Discovery Rate correction, considering three main comparisons: the difference between post-treatment and baseline values for the active and distractor treatments independently, and the comparison of this difference (post-treatment versus baseline) between the two treatments. Additional outcome measures were not corrected for multiple comparisons. Effect sizes were measured using Cohen’s d with scores between 0.2 and 0.49 considered to be small effects, between 0.5 0.79 medium effects and ≥ 0.8 large effects [12]. In accordance with previous studies on changes in chronic pain intensity using an 11-point scale (0 no pain to 10 pain as bad as possible) [60,61], we considered a change of two points or more (∼30% change) on the primary outcome measures to be clinically significant. Interim analyses were not conducted as not permitted by the pre-registered protocol, but descriptive patterns in the raw data were examined every 6 months. For the primary outcome measure, participants who had a missing score at one of the times points were excluded from the analyses (post-treatment - baseline [active treatment: n = 1]; 1-week follow-up minus baseline [distractor treatment: n = 1]; 8-week follow-up minus baseline [active treatment: n = 3; distractor treatment: n =1]). All participants (n=36) were included in the analysis of the secondary outcome measure.

Data, scripts and testing materials are available on OSF (https://osf.io/xsd4e).

## 3. RESULTS

### 3.1. Primary outcome measures: Average pain ratings at baseline versus post-treatment

Table 1 shows scores on one of the primary outcome measures, present pain intensity ratings from the McGill visual-analog scale, at baseline, post-treatment, and at one-and eight-week follow-up.^1^ Permutation analyses showed that reduction in pain severity post-treatment was statistically significant and associated with a moderate effect size for the active (M =-1.42, SE=0.52, p=0.002, q = 0.006, Cohen’s d=0.60) but not for the distractor treatment (M =-.29, SE= 0.73, p=0.34 q=0.34, d=0.10) (See Figure 3). The difference between the two treatments did not reach significance (p=0.10, q=0.15).

**Figure 3:**
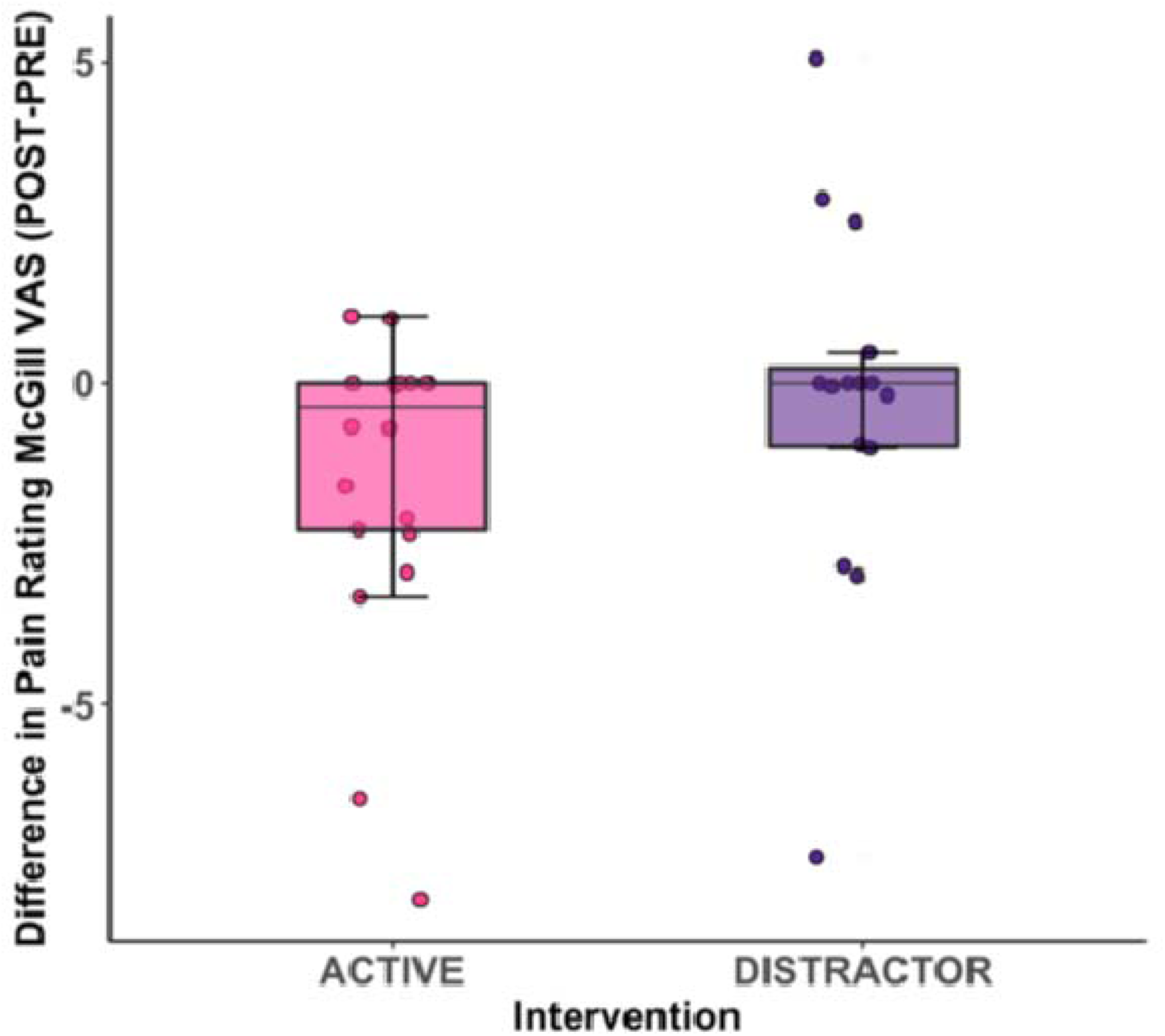
Box plots of primary outcome measure (differences in pain intensity rating of the McGill Visual Analogue Scale post-treatment minus baseline) for the active and distractor treatment groups. Larger negative numbers indicate larger treatment effects. The error bars indicate the upper and lower data points within a 1.5 Interquartile Range; the box represents the upper, middle and lower quartile.

**Table 1:**
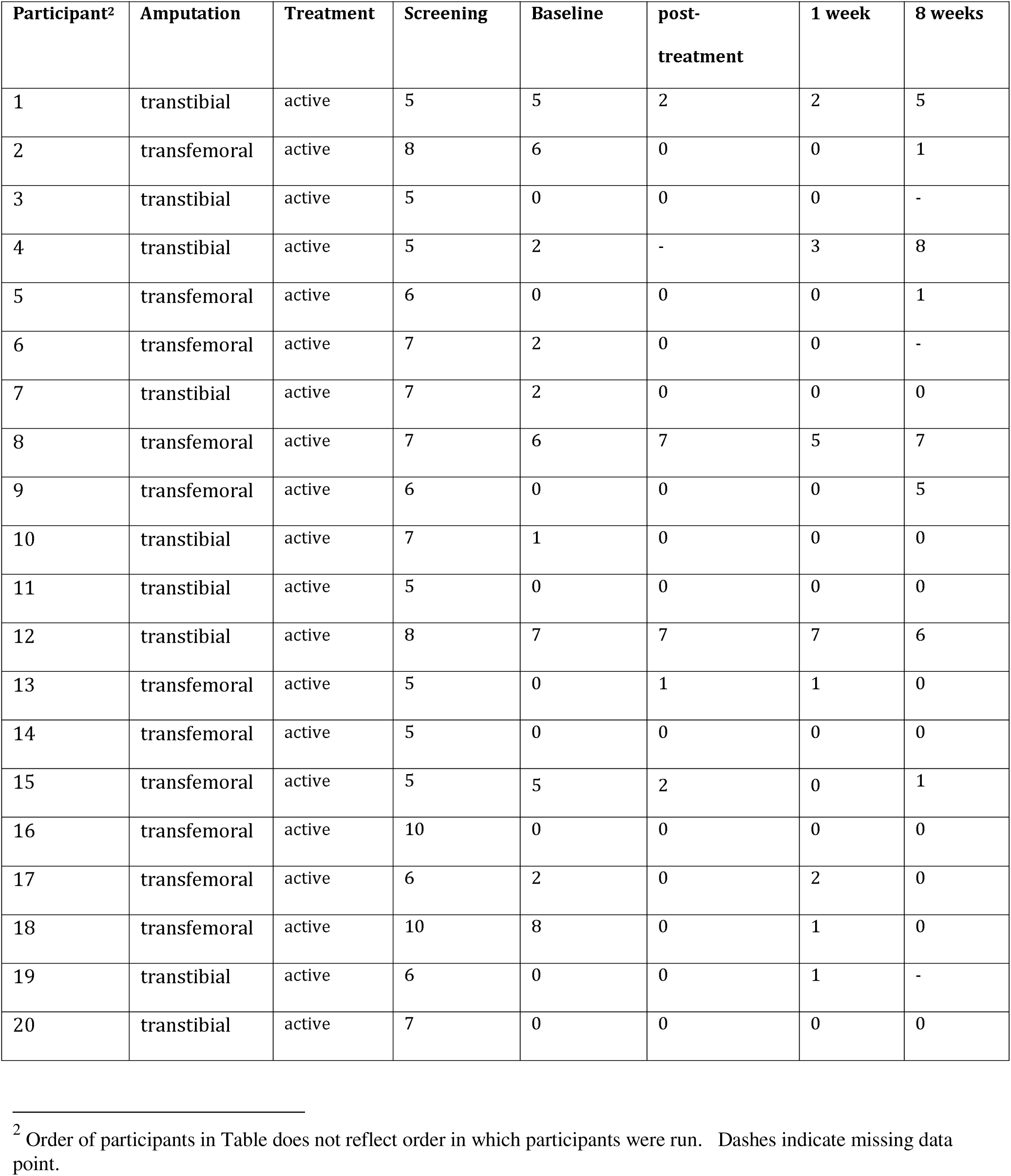
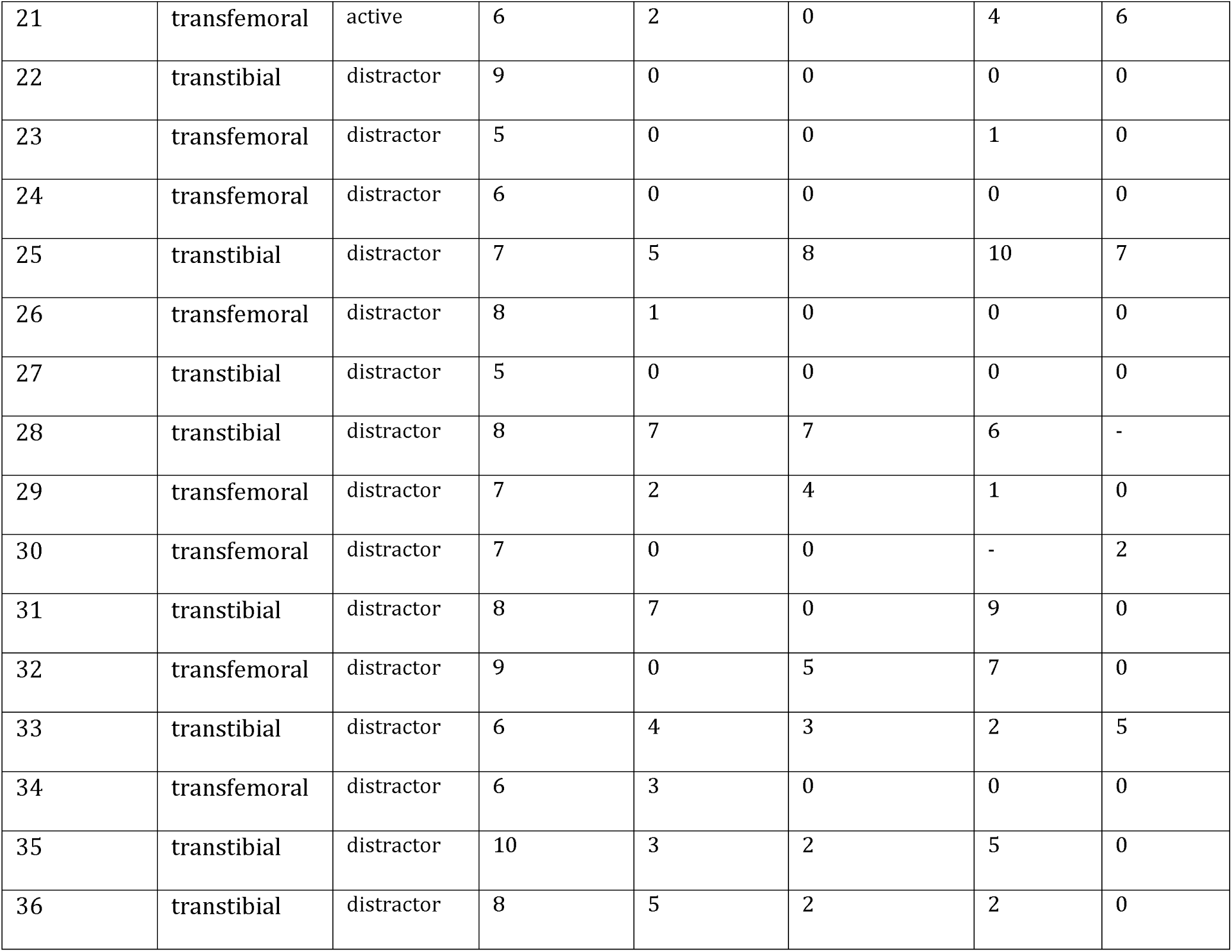
McGill Visual Analogue Scale Scores for pain intensity at baseline, post-treatment, one-week follow-up, and eight-week follow-up for all participants.

Collapsing across study arms, 11 (31.4%) of the 35 participants who have data at both baseline and immediate post-treatment showed a numerical improvement in pain of at least two points [60,61]. Eight participants in the active group (40%) and 3 (20%) in the distractor group showed clinically significant pain reduction.

In terms of the level at which the leg was amputated, a similar clinically significant improvement of two points or more at baseline versus immediate post-treatment timepoints was observed for transfemoral and transtibial amputees (see Table 2). Similar patterns of clinically significant improvement were also observed when the two treatments were examined separately for effects of level of amputation.

**Table 2:**

| **Overall sample** | Active (n=20) | Distractor (n=15) | Chi2 |
| --- | --- | --- | --- |
| Improvement ($\geq 2$ points) | 8 (40%) | 3 (20%) | $\chi^2 = 1.59$,<br>$p = .207$, $V = 0.21$ |
| No clinically meaningful improvement | 12 (60%) no change | 12 (80 %):<br>-9 no change<br>-3 worsening $\geq 2$ points | |
| **Overall sample** | Transfemoral (n=19) | Transtibial (n=16) |  |
| Improvement ($\geq 2$ points) | 7 (37%) | 4 (25%) | $\chi^2 = 0.57$,<br>$p = .452$, $V = 0.13$ |
| Active | 6 (of 12; 50%) | 2 (of 8; 25%) | $\chi^2 = 1.25$,<br>$p = 0.26$, $V = 0.25$ |
| distractor | 1 (of 7; 14%) | 2 (of 8; 25%) | $\chi^2 = 0.26$,<br>$p = 0.60$, $V = 0.13$ |

A similar pattern was observed when examining the difference (post treatment-baseline) in pain severity based on the qualitative pain questions of the McGill Pain questionnaire. Permutation analyses showed that the decrease in pain severity was marginally significant and associated with a moderate effect size for the active (M=-6.45, SE= 2.83, p=0.017, q =0.051, d=0.51) but not for the distractor treatment (M=-2.46, SE= 2.15, p=0.15, q=0.15, d=0.19). The difference between the two treatments was not statistically significant (p=0.16, q=0.15).

To address the question of persistence of treatment effects one-week post-treatment, we compared the McGill visual analog scale present pain ratings obtained at baseline with the one-week follow-up session (see Table 1 for participants’ data). For the active treatment, we observed a persistent reduction in pain associated with a moderate effect size (M= - 1.24, SE=0.55, p=0.016, q = 0.048, Cohen’s d=0.50). No significant effects were observed for the distractor treatment (M=-0.34, SE=0.70, p=0.67, q=0.67, d=-0.12). There was a numerical difference between treatments in favor of the active treatment that did not survive correction for multiple comparisons (p=0.04, q =0.06).

We also examined persistence of reduction in pain severity pre-treatment versus eight-week follow-up with the McGill visual analog scale. No significant differences were observed between baseline and eight-week follow-up for either the active (M=0.76, SE=0.76, p=0.17, q = 0.25, d=0.24) or distractor treatment (M=1.11, SE=0.71, p=0.07, q =0.21, d=0.41), nor was there a difference between treatments (p=0.63, q=0.63).

The Pain Rating Index from the McGill questionnaire, which assesses qualitative characteristics of pain, showed significant, moderate effect sizes in the comparisons between baseline versus one-week follow-up for both the active treatment (M=-6.2, SE=2.38, p=0.009, q=0.03, d=0.58) and distractor treatment (M=-5.64, SE=2.59, p=0.02, q=0.03, d=0.58). No significant differences were observed between the two treatments (p=0.43, q=0.43).

The difference in the Pain Rating Index at baseline versus eight-week follow-up was significant and associated with a moderate effect size for the active treatment (M=-8.11, SE=2.90, p=0.0004, q=0.012, d=0.67) but not for the distractor treatment (M=-3.86, SE=2.49, p=0.07, q=0.1, d=0.41). The difference between the two treatments was not significant (p=0.13, q=0.13).

### 3.2. Secondary outcome measure: Average pain intensity since the last session

Figure 4 shows the average pain rating since the last session (a measure modified from the Brief Pain Inventory [10]) for the data collected at the beginning of each treatment session.

**Figure 4:**
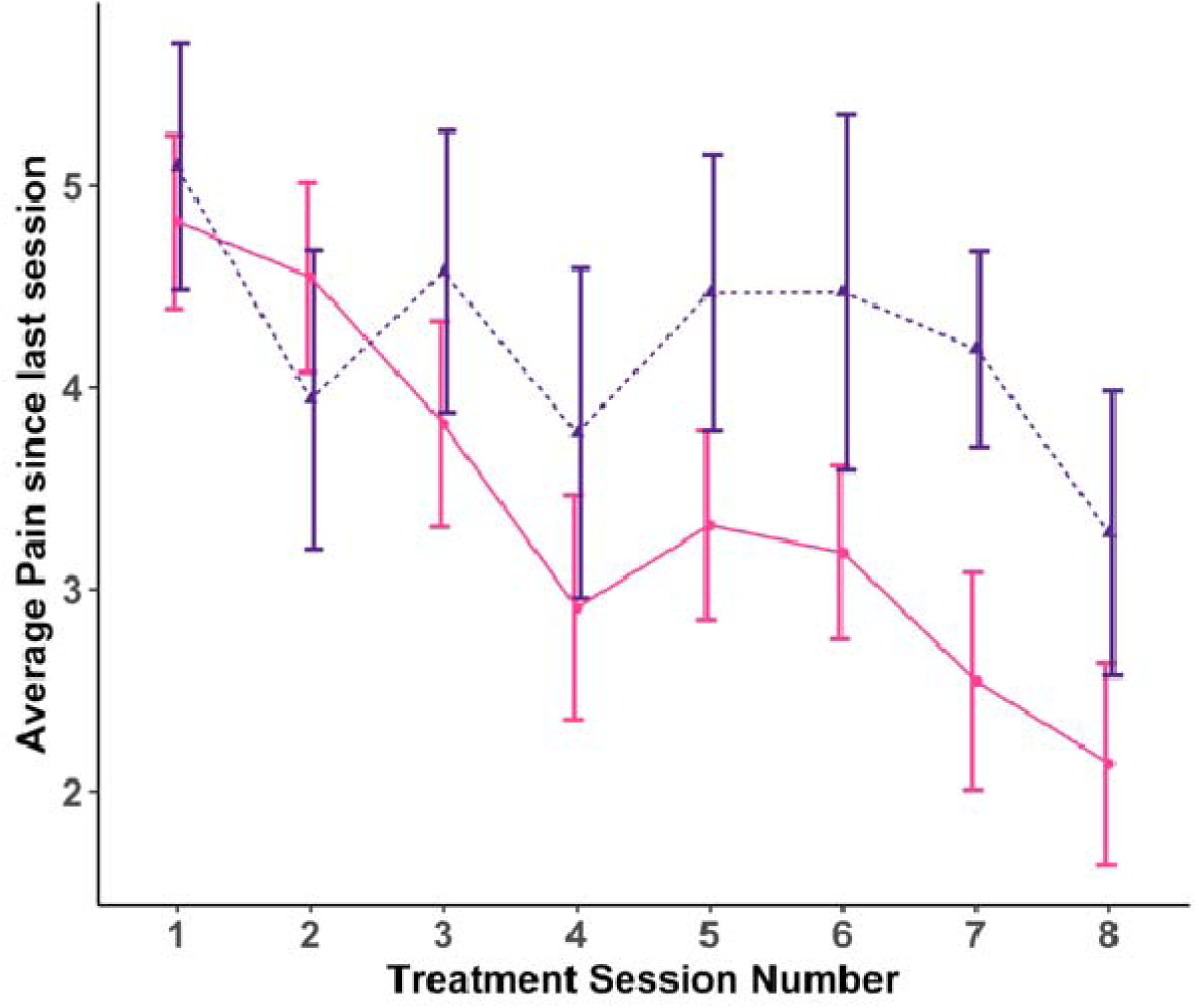
Plots of secondary outcome measure: Ratings of average pain intensity since the last session for active (pink) and distractor (purple) treatment groups across sessions.

We observed a progressive decrease in pain levels across sessions for both active and distractor conditions. Permutation analyses conducted on the difference in first-versus last-session ratings indicated that there was a significant and large effect for the active treatment (M=-2.68, SE=0.44, p<0.001, q=0.001, d=1.27) and a significant but moderate effect for the distractor treatment (M=-1.81, SE=0.83, p=0.026; q=0.039, d=0.56). The difference between treatments did not reach significance (p=0.16, q=0.16).

### 3.3. VR usability and presence assessments

The Technology Acceptance Scale administered at baseline showed that participants in the two arms of the study were similarly highly accepting of technology (Active Treatment M=24, SD=9.5; Distractor Treatment M=29, SD=7.5). Data from the Simulator Sickness Questionnaire collected after the first and final treatment indicated similar low levels of cybersickness for the active (1st session M=0.17, SD=0.20; 8th session M=0.18, SD=0.20; p=0.74) and distractor (1st session M=0.07, SD=0.11; 8th session M=0.16, SD=0.24; p=0.17) treatments. Similar results were observed in the two testing sessions for the Slater-Usoh Steed Presence Questionnaire for both active (1st session M=4.79, SD=1.24; 8th session M=4.65, SD=1.53; p=0.59) and distractor (1st session M=4.77, SD=1.16; 8th session M=4.72, SD=1.10; p=0.43). Participants indicated a similar strong feeling of presence (p=0.29) and low level of cybersickness (p=0.87) in the active and distractor treatments. Participants reported high usability of both the active (M=34.4, SD=14.6) and distractor (M=30.6, SD=3.3) treatments, with no significant difference between the two treatments (p=0.26). Participants’ satisfaction was high for both active (M=8.08, SD=3.08) and distractor (M=8.94, SD=1.48) treatments (p=0.31).

### 3.4. Additional test battery measures

Participants also completed several additional questionnaires to assess pain at its worst, interference of pain in daily life, depression, anxiety, and quality of sleep. Table 3 shows the mean, standard error, and number of observations for these additional questionnaires presented at baseline, post-treatment, one-and eight-week follow-up sessions. Permutation analyses (uncorrected) were used for these exploratory comparisons.

**Table 3:** Descriptive statistics for the battery of questionnaires at baseline, post-treatment, and at 1-and 8-week follow-up for the active and distractor treatments.

|  |  | ACTIVE |  |  |  |  | DISTRACTOR |  |  |  |  |
| --- | --- | --- | --- | --- | --- | --- | --- | --- | --- | --- | --- |
|  |  | Baseline | Post treatment | 1 week follow up | 8 week follow up | Difference Post-Baseline treatment | Baseline | Post treatment | 1 week follow up | 8 week follow up | Difference Post-Baseline treatment |
| VAS PLP worst | *mean* | 7.95 | 5.77* | 5.36* | 5.52*# | -2.18 | 7.77 | 6.94 | 6.68 | 6.94*# | -0.83 |
|  | *se* | 0.09 | 0.14 | 0.10 | 0.14 | 0.67 | 0.09 | 0.17 | 0.23 | 0.12 | 0.55 |
|  | *n* | 21 | 20 | 21 | 18 | 20 | 15 | 15 | 14 | 14 | 15 |
| PLP frequency | *mean* | 3.05 | 4.30 | 3.95 | 4.44 | 1.30 | 3.20 | 4.13 | 4.43 | 4.86 | 0.93 |
|  | *se* | 0.06 | 0.10 | 0.10 | 0.13 | 0.08 | 0.07 | 0.11 | 0.15 | 0.15 | 0.12 |
|  | *n* | 21 | 20 | 21 | 18 | 20 | 15 | 15 | 14 | 14 | 15 |
| PLP interference | *mean* | 22.63 | 12.24* | 12.39* | 15.25 | -9.02 | 22.38 | 22.56 | 14.94 | 18.46 | -0.29 |
|  | *se* | 0.98 | 0.78 | 0.70 | 0.85 | 1.09 | 1.16 | 1.55 | 1.38 | 1.17 | 1.47 |
|  | *n* | 21 | 20 | 21 | 18 | 20 | 15 | 15 | 14 | 14 | 15 |
| FAI | *mean* | 21.62 | 22.65 | 21.86 | 22.89 | 0.70 | 22.60 | 21.20 | 22.36 | 21.57 | -1.47 |
|  | *se* | 0.36 | 0.37 | 0.42 | 0.48 | 0.24 | 0.68 | 0.66 | 0.65 | 0.72 | 0.33 |
|  | *n* | 21 | 20 | 21 | 18 | 20 | 15 | 15 | 14 | 14 | 15 |
| **SF-12 physical** | *mean* | 40.52 | 41.53 | 44.92 | 43.52 | -0.53 | 38.75 | 41.20 | 38.50 | 43.79 | 4.04 |
|  | *se* | 0.74 | 0.64 | 0.56 | 0.80 | 0.55 | 0.85 | 1.25 | 1.47 | 0.65 | 1.16 |
|  | *n* | 21 | 20 | 21 | 18 | 20 | 15 | 15 | 14 | 14 | 15 |
| **SF-12 mental** | *mean* | 40.52 | 55.01 | 52.87 | 50.63 | 2.30 | 45.43 | 53.85 | 50.65 | 49.16 | 7.90 |
|  | *se* | 0.74 | 0.49 | 0.79 | 0.79 | 0.95 | 1.40 | 0.83 | 0.82 | 1.44 | 1.25 |
|  | *n* | 21 | 20 | 21 | 18 | 20 | 15 | 15 | 14 | 14 | 15 |
| **HADS - anxiety** | *mean* | 5.52 | 4.65 | 4.62 | 5.33 | -0.95 | 5.93 | 5.00 | 5.64 | 4.07* | -0.53 |
|  | *se* | 0.18 | 0.17 | 0.18 | 0.22 | 0.13 | 0.28 | 0.33 | 0.35 | 0.29 | 0.22 |
|  | *n* | 21 | 20 | 21 | 18 | 20 | 15 | 15 | 14 | 14 | 15 |
| **HADS - depression** | *mean* | 4.57 | 4.30 | 3.57 | 3.89 | -0.20 | 4.13 | 4.67 | 3.86 | 3.93 | 0.53 |
|  | *se* | 0.14 | 0.17 | 0.16 | 0.20 | 0.11 | 0.24 | 0.27 | 0.30 | 0.24 | 0.18 |
|  | *n* | 21 | 20 | 21 | 18 | 20 | 15 | 15 | 14 | 14 | 15 |
| **Pain Catastrophizing** | *mean* | 19.95 | 19.15 | 18.67 | 18.28 | -1.60 | 16.20 | 14.47 | 12.71 | 10.86 | -0.40 |
|  | *se* | 0.57 | 0.63 | 0.65 | 0.66 | 0.51 | 0.88 | 0.93 | 0.88 | 0.81 | 0.82 |
|  | *n* | 21 | 20 | 21 | 18 | 20 | 15 | 15 | 14 | 14 | 15 |
| **Insomnia** | *mean* | 9.81 | 6.95 | 8.29 | 8.50 | -2.20 | 10.93 | 9.33 | 8.93 | 10.00 | -1.67 |
|  | *se* | 0.41 | 0.38 | 0.37 | 0.46 | 0.40 | 0.38 | 0.61 | 0.57 | 0.51 | 0.53 |
|  | *n* | 21 | 20 | 21 | 18 | 20 | 15 | 15 | 14 | 14 | 15 |

\*p < .05 for treatment effect, #p < .05 for comparison between treatments;
VAS: visual analogue scale; FAI: Frenchay Activities Index; SF-12: Twelve-Item Short Form Health Survey; HADS: Hospital

The visual analogue scale of the Limb Deficiency and Phantom Limb Pain questionnaire, which assesses pain at its worst, showed a moderate effect size for decrease in PLP in the post-treatment-versus baseline comparison for the active (M=-2.18, SE=0.67, p=0.002, d=0.72) but not for the distractor treatment (M=-0.83, SE=0.55, p=0.07, d=0.39); the difference between the two treatments did not reach statistical significance (p=0.07).

Similar improvements in PLP at its worst were observed comparing baseline and 1-week follow-up; and baseline and 8-week follow-up for the active treatment (both p ≤ 0.001 with moderate effect sizes). For the distractor treatment, we observed an improvement in’pain at its worst’ when comparing baseline-with 8-week follow-up (p=0.003), but not when comparing baseline-with 1-week follow-up (p=0.09). The two treatments differed in ‘pain at its worst’ when comparing the baseline-with the 8-week follow-up session, with the active treatment (M=-2.65, SE=0.72) showing a statistically larger improvement than the distractor treatment (M=0.85, SE=0.27) (p=0.02), while the difference did not reach significance in the comparison of baseline-with the 1-week follow-up (p=0.09).

Similar results were observed for the pain interference in daily life questionnaire. For the active treatment, there was a significant difference between the baseline and post-treatment values, with a moderate effect size (M=-9.37, SE=4.82, p=0.03, d=0.43). There was a statistically significant effect with a moderate effect size for the difference between baseline and 1-week follow-up (M=-10.24, SE=4.10, p=0.006, d=0.54). These differences were not significant for the distractor treatment (ps>0.09). The differences between the two treatments were not significant in any of the comparisons (ps>0.30).

No additional significant effects were observed, except for a significant effect of the distractor treatment for the anxiety score of the Hospital Anxiety and Depression Scale (HADS) for the difference between baseline and 8-week follow-up, with a moderate effect size (M=-1.78, SE=0.83, p=0.03, d=0.57).

### 3.5. Predictors of changes in pain scores

We conducted an exploratory linear regression analysis with data from both the active and distractor treatments together to assess whether time since amputation (in days), cause of amputation (1. Surgical, 2. Traumatic), sense of agency over the phantom (presence of agency, absence of agency), or perceived size of the phantom limb (1. Normal, 2. Enlarged, 3. Shrunken) predicted changes in PLP measured with the McGill VAS baseline-versus post-treatment. The results of the regression indicated that the model was significant, F(2, 28) = 2.80, p = 0.04, R^2^=0.27. Significant predictors of changes in PLP were time since amputation (t=-2.70, p=0.011) and cause of amputation (t= 2.1, p=0.042). These data suggest that VR treatment may be most beneficial shortly after amputation and when the amputation is traumatic rather than surgical.

### 3.6. Adverse events

There were three adverse events: two were minor, expected, and related to the study, and one was severe, unexpected, and unrelated to the study. The two minor adverse events consisted of cybersickness experienced during the distractor treatment (n = 1) and the active treatment (n = 1). The severe adverse event was the death of a participant enrolled in the distractor treatment arm. The participant’s death did not occur during treatment and was caused by a health condition unrelated to the study.

## 6. Discussion

This registered clinical trial comparing the efficacy of two VR interventions for phantom limb pain (PLP) showed several important effects. First, our primary outcome measures, ratings of pain severity and qualitative characteristics of pain (Pain Rating Index) from the McGill Pain Questionnaire at baseline versus after the completion of 8 sessions of treatment, showed significant improvements and moderate effect sizes for the active treatment but not for the distractor treatment. These effects of the active treatment were persistent at 1-week follow-up for both pain severity and quality, and at 8-week follow-up for pain quality. The distractor treatment showed significant positive effects at 1-week for pain quality. Although the active treatment showed significant and long-lasting effects, superiority of this treatment was not established.

Our secondary outcome measure, average pain intensity since the last session as measured across testing sessions, showed significant positive effects for both active and distractor treatments. Although the effect size was numerically larger for the active treatment, the difference between the two treatments was not statistically significant. We also assessed responses on a number of additional measures. Pain at its worst and pain interference in daily life were significantly reduced, with moderate effect sizes, after the active but not distractor treatment. Moreover, some effects persisted at 1-and/or 8-weeks post-treatment, and reduction in “pain at its worst” was significantly more favorable for the active than distractor treatment when comparing baseline to the 8-week follow-up. While direct comparisons between treatments did not reach statistical significance for other pain outcomes, the results as a whole suggest that the benefits of the active treatment extended beyond pain perception *per se* and translated into meaningful improvements in everyday functioning. Finally, participants in the present study only reported mild and similar cybersickness during both treatments, and both approaches were viewed as highly usable. Overall, treatment compliance and participant satisfaction were high for both treatments. Both treatments were safe and generally well tolerated.

Virtual reality (VR) interventions for phantom limb pain (PLP) have been shown to have positive effects on pain reduction [2,35,39], and the present study confirms and extends this evidence. A broad body of literature indicates that VR exerts a general analgesic effect during exposure, likely through distraction and emotional engagement. Indeed, VR experiences have been shown to reduce pain perception during a range of painful medical procedures, including venipuncture, childbirth, burn dressing changes, and chemotherapy (see [25,52]). Furthermore, consistent with the present data, interactive VR has been shown to produce greater analgesic effects than passive VR experiences [47].

The observed differences in “pain at it’s worst” following the two treatments invite consideration of the possible mechanisms underlying the two VR approaches. Although both treatments were immersive and engaging, they were based on distinct therapeutic principles. The distractor treatment relied primarily on attentional analgesia, whereby an entertaining VR experience diverts attention away from pain and temporarily reduces pain perception. In contrast, the active VR treatment allowed participants to view a virtual rendering of their amputated limb as intact and to interact with the environment using a controller placed on the stump. This configuration enabled participants to control and embody the virtual limb, likely fostering a sense of agency over the VR leg. Such embodiment-related processes may have exerted a more specific influence on the phantom limb representation, thereby contributing to longer-lasting reduction in pain at its worst. This speculative interpretation is consistent with recent work showing superior outcomes when active VR training is combined with traditional rehabilitation, compared to rehabilitation alone [1].

Importantly, the robustness of these findings is supported by the methodological rigor of the study design. Our conservative approach involved comparing our active VR treatment with another well-established, efficacious VR-based pain intervention rather than with a placebo condition. This choice may have reduced the observed contrast between treatment arms. Stratified randomization accounted for several factors known to be associated with PLP, including pain severity, level of amputation, telescoping, and prosthesis use.

Telescoping—a phenomenon in which the phantom limb is perceived as shortened, with the foot felt closer to the stump [16,20]—has been positively associated with PLP severity [17,22,40], and has been shown to reduce treatment efficacy in mirror box interventions [18]. In addition, PLP influences prosthesis use, as individuals without PLP are more likely to use their prosthesis consistently [43]. By incorporating these variables into the randomization process, the present study ensured a balanced comparison between treatments, supporting confidence in the observed efficacy of the active intervention.

At the individual level, response to treatment varied considerably, highlighting the heterogeneous nature of PLP [15]. In the total sample, 31.4% of participants showed a clinically-significant reduction in pain intensity at the end of treatment (40% in the active and 20% in the distractor group). When compared with our previous study [2], which reported pain improvement in 85% of participants, the present results appear more modest. However, participants in the earlier study all reported moderate-to-severe pain at baseline, whereas the present sample included several individuals with little or no baseline pain. This may reflect differences between our inclusion criterion, which focused on average pain over the preceding month, and our baseline measures focusing on current pain. Furthermore, in our previous work we considered treatment to be beneficial with any reduction in pain intensity while in the present study we used the more conservative but clinically-relevant criterion of approximately 30% pain reduction [60,61].

The present study also extends previous work by demonstrating comparable efficacy across levels of amputation. Numerical pain improvement was observed in similar proportions among individuals with transfemoral and transtibial amputations. These findings are particularly relevant because most VR-based PLP studies have focused primarily on transtibial populations [2,13], while studies including participants with both levels of amputations have either been case reports or dominated by transtibial samples [1,19]. To our knowledge, this is the first study to employ the same VR apparatus in a relatively large and balanced sample of transfemoral and transtibial amputees, demonstrating similar applicability and therapeutic benefit across these populations.

Given the variability in individual responses, an important question concerns which patients may benefit most from VR-based interventions. To address this issue, we conducted an exploratory regression analysis examining predictors of PLP change. Results suggest that treatment may be more effective when delivered closer to the time of amputation and in cases of traumatic rather than surgical amputation. Although these findings should be interpreted cautiously due to the limited sample size, they are consistent with previous evidence indicating that PLP severity is influenced by time since amputation [22] and that PLP is more prevalent in traumatic amputees [45]. These observations suggest that VR interventions may be particularly effective when applied early, during the acute phase of PLP, potentially preventing chronic pain development.

Several considerations also emerge regarding optimization of treatment parameters. In the present study, VR sessions lasted approximately one hour and were delivered twice weekly over four weeks. This duration was chosen to balance feasibility with sufficient treatment intensity and aligns with previous work demonstrating efficacy using similar protocols [2,3,11]. However, positive effects of treatment for PLP have also been reported following shorter (10–30 minutes) or longer (∼2 hours) sessions [46,54]. Future studies should examine dose–response relationships, including the minimum effective session duration, the persistence of analgesic effects following each session, and the relationship between game-play time and pain reduction.

The number of treatment sessions also warrants further investigation. While some studies have demonstrated pain relief following a single VR session [46], longer-lasting effects typically require repeated exposure [54]. In the present study, eight sessions were selected to minimize participant dropout, as extended in-person treatment commitments are often impractical. Notably, some participants continued to report pain after completing the protocol, and several showed a gradual reduction in pain that had not reached asymptote at the conclusion of treatment, suggesting that additional sessions may have produced further benefit in this group. This raises the possibility that extending treatment or tailoring session numbers to individual responses could enhance outcomes.

Although the effects of the active VR treatment on pain quality and intensity of pain at its worst persisted for up to eight weeks, longer-term efficacy remains unknown. Future studies should assess maintenance of treatment effects beyond this timeframe and determine whether pain recurs. In such cases, occasional booster sessions may be an effective strategy to reinforce and prolong therapeutic benefits.

A limitation of the present study, noted above, was the mismatch between the study inclusion criteria, which focused on average pain scores over the prior month, and the primary outcome measures, which assessed changes in current pain relative to participants’ baseline pain. Given the variability of PLP, several participants reported no pain during the baseline assessment, resulting in no possibility of observing improvement in pain scores across time points. This limitation may have led to an underestimation of the treatment effects

In conclusion, the present study confirms VR as an efficacious and safe intervention for PLP in both transtibial and transfemoral amputees. Although superiority of one treatment over another was not established for most measures, the active VR treatment—requiring participants to view and control an intact virtual limb using their amputated limb—yielded consistent, functionally meaningful, and long-lasting benefits. These findings support active VR as a valuable non-pharmacological tool for PLP management. When viewed in the context of the treatment’s high ratings for usability and tolerability, the data underscore the feasibility and clinical promise of active VR interventions for PLP.

## Data Availability

All data and program codes are available at the following link https://osf.io/xsd4e.

## Acknowledgments

The authors have no conflicts of interest to declare. All data and program codes are available at the following link https://osf.io/xsd4e. Generative AI was used to check grammar. The study was funded by the National Institutes of Health (NIH) (R01 HD104158).

## Footnotes

1 Note that participants were enrolled in the study based on average pain scores > 4 over the prior month, whereas the primary outcome measures assessed current pain at the time of the baseline assessment. This resulted in inclusion of some participants who had no current pain at baseline.

## Notes

### Competing Interest Statement

The authors have declared no competing interest.

### Clinical Trial

NCT05296265

### Author Declarations

This study was conducted with full approval of the Institutional Review Board of the Office of Human Research Protection, Sidney Kimmel Medical College, Thomas Jefferson University (IRB # AEHN2022-775).

### Summary of Updates

We have added a table which summarize part of the results

